# Efficacy and safety of neoadjuvant stereotactic body radiation therapy plus dalpiciclib and exemestane for hormone receptor-positive, HER2-negative breast cancer: A prospective pilot study

**DOI:** 10.1101/2024.08.31.24312890

**Authors:** Yu Zhang, Shuo Cao, Nan Niu, Huilian Shan, Jinqi Xue, Guanglei Chen, Yongqing Xu, Jianqiao Yin, Chao Liu, Lisha Sun, Xiaofan Jiang, Meiyue Tang, Qianshi Xu, Mingxuan Jia, Xu Zhang, Zhenyong Zhang, Qingfu Zhang, Jianfei Wang, Ailin Li, Yongliang Yang, Caigang Liu

**Author notes:** Corresponding author: Caigang Liu, MD, PhD, Department of Oncology, Shengjing Hospital of China Medical University, Shenyang 110004, China. **Authors’ contributions:** Y. Zhang, Data curation, formal analysis, methodology. S. Cao, Data curation, formal analysis, methodology, writing–original draft. N. Niu, Validation, data curation, formal analysis, methodology, writing–review and editing. H. Shan, Data curation, methodology. J. Xue, Data curation, methodology. G. Chen, Data curation, methodology. Y. Xu, Methodology. J. Yin, Methodology. C. Liu, Formal analysis, methodology. L. Sun, Methodology. X. Jiang, Writing-original draft. M. Tang, Data curation. Q. Xu, Formal analysis. M. Jia, Methodology. X. Zhang, Methodology. Z. Zhang, Methodology. Q. Zhang, Formal analysis. J. Wang, Methodology. A. Li, Conceptualization, methodology. Y. Yang, Methodology, writing–review and editing. Caigang Liu, Conceptualization, supervision, project administration, writing–review and editing. Yu Zhang, Shuo Cao, Nan Niu, Huilian Shan and Jinqi Xue contributed equally to this work. **Clinical trial number: NCT05132790**. **Impact statement: Neoadjuvant SBRT with dalpiciclib and exemestane resulted in a 16.7% RCB 0-I rate and a 91.7% ORR, with acceptable toxicities in HR-positive, HER2-negative breast cancer patients.**.

## Abstract

**Background:** Both neoadjuvant chemotherapy and endocrine therapy only result in trivial pathological complete response rates and moderate objective response rates (ORR) in hormone receptor (HR)-positive, human epidermal growth factor receptor-2 (HER2)-negative breast cancer, more promising alternatives are urgently needed. With proven synergistic effect of cyclin dependent kinase 4/6 (CDK4/6) inhibitor and radiotherapy in preclinical studies, this pilot study aimed to explore the efficacy and safety of neoadjuvant stereotactic body radiation therapy (SBRT) followed by dalpiciclib and exemestane in HR-positive, HER2-negative breast cancer.

**Methods:** This was a single-arm, non-controlled prospective pilot study. Treatment-naïve patients with unilateral HR-positive, HER2-negative breast cancer received neoadjuvant radiotherapy (24Gy/3F) followed by dalpiciclib and exemestane for 6 cycles. The primary endpoint was the proportion of patients with residual cancer burden (RCB) score of 0-I. Key secondary endpoints included ORR, breast-conservation rate, biomarker analysis, and safety.

**Results:** All 12 enrolled patients completed the study treatment and surgery. Two (16.7%) of them achieved the RCB 0-I with the ORR of 91.7% (11/12). Analyses of tumor specimens showed significant increase of infiltrating T cells rather than alteration of PD-L1 positive immune cells. The most common grade 3 adverse events (AEs) were neutropenia (66.7%) and leukopenia (25.0%), but no grade 4-5 AE or death occurred.

**Conclusions:** Our results suggested neoadjuvant SBRT followed by dalpiciclib and exemestane are effective and tolerable, and provides novel insights for the neoadjuvant treatment of HR+/HER2-breast cancer, which may be considered as a feasible option for patients with HR-positive, HER2-negative breast cancer.

## Introduction

Hormone receptor (HR)-positive, human epidermal growth factor receptor 2 (HER2)-negative breast cancer is the most prevalent subtype of breast cancer, which accounts for approximately 70% of all breast cancer (Parker et al., 2009). Neoadjuvant chemotherapy (NCT) is recommended as the first-choice for patients with HR-positive, HER2-negative breast cancer, who are candidates for preoperative therapy, according to National Comprehensive Cancer Network (NCCN) guidelines(Gradishar et al., 2022). Neoadjuvant endocrine therapy (NET) is an acceptable, less toxic alternative of NCT. However, less than 10% of patients with HR-positive, HER2-negative breast cancer can achieve pathological complete response (pCR), and only 60% of patients respond to NCT or NET (Sella et al., 2021). Hence, more promising alternatives are urgently needed.

Cyclin dependent kinase 4/6 (CDK4/6) inhibitors are breakthroughs in the treatment of HR-positive, HER2-negative breast cancer, and have a remarkable efficacy in the advanced setting when combined with endocrine therapy (ET) (Finn et al., 2015; Goetz et al., 2017; Hortobagyi et al., 2016). Presently, abemaciclib has been approved for high-risk, early HR-positive breast cancer plus ET as adjuvant treatment and its application is being explored in neoadjuvant setting (neoMONARCH and NCT04293393). However, therapies with combination of CDK4/6 inhibitors and NET only achieve a pCR rate of less than 5% (Hurvitz et al., 2020; Johnston et al., 2019).In the PALLET study, objective response rate (ORR) was not statistically significantly different between the combinational therapy and monotherapy (54.3% vs 49.5%, P = 0.20) (Johnston et al., 2019). Likewise, in the FELINE study, there was no statistical difference in clinical, mammographic, ultrasound or magnetic resonance imaging (MRI) response between letrozole alone and combination of letrozole with a CDK4/6 inhibitor(Khan et al., 2020). Thus, it is reasonable to explore a novel neoadjuvant therapy for HR-positive, HER2-negative breast cancer.

Radiotherapy is one of the most important local methods to control malignancy by breaking DNA in tumor cells (Cox & Swanson, 2013). Stereotactic body radiation therapy (SBRT) is a novel and relatively safe radiotherapy technique and delivers high doses of radiation in a small number of fractions to the tumor lesion while minimizing radiation exposure to the surrounding tissues(Chmura et al., 2021). Currently, neoadjuvant radiotherapy can be applicable for inoperable breast cancer to allow an effective and sometimes more conservative surgery(Cardoso et al., 2018). Preclinical studies have indicated that CDK4/6 inhibitors can sensitize breast cancer cells to radiotherapy, possibly by inhibition of DNA damage repair response, enhancement of apoptosis and blockage of cell cycle progression, and inhibition of intra-tumor cellular metabolism and angiogenesis (Han et al., 2013; A. M. Pesch et al., 2020; G. Petroni et al., 2021). A preclinical study has explored different schedules of combination of radiotherapy and CDK4/6 inhibitors in human and mouse model and demonstrated that radiotherapy followed by CDK4/6 inhibitors can enhance antineoplastic effects, compared with their monotherapy alone or CDK4/6 inhibitors concurrent with or followed by radiotherapy (G. Petroni et al., 2021). Moreover, the combination of CDK4/6 inhibitors and radiotherapy can lead to a satisfying overall disease control and safety based on several retrospective studies in metastatic breast cancer (Bosacki et al., 2021). Nevertheless, the efficacy and safety of this regimen in neoadjuvant treatment of early or locally advanced HR-positive, HER2-negative breast cancer are unknown. We have conducted a prospective pilot study to determine the efficacy and safety of neoadjuvant radiotherapy followed by dalpiciclib and exemestane in patients with newly-diagnosed early or locally advanced HR-positive, HER2-negative breast cancer. Dalpiciclib (SHR6390) is a potent selective inhibitor of CDK4/6 and exerts synergistic anti-tumor effect when combined with endocrine therapy for advanced luminal breast cancer in the DAWNA-1 trials (Xu et al., 2021).

## Methods

### Study design

This single-arm, non-controlled prospective pilot study in women with HR-positive, HER2-negative early or locally advanced breast cancer was registered with ClinicalTrials.gov (NCT05132790).

The study was approved by the Institutional Review Board and Ethics Committee of Shengjing Hospital of China Medical University, according to the Declaration of Helsinki and Good Clinical Practice guidelines. Written informed consent was obtained from each patient.

### Participants

Eligible patients included pre- and post-menopausal women aged between 18 and 75 years, with histologically confirmed HR-positive, HER2-negative (0 or 1+ by immunohistochemistry [IHC], or 2+ by IHC with negative results of fluorescence in situ hybridization), early or locally advanced invasive breast cancer with the lesion of ≥2cm by MRI. Other main inclusion criteria were Eastern Cooperative Oncology Group performance status 0-1, adequate marrow, hepatic and renal function. The key exclusion criteria included stage IV tumors, inflammatory breast cancer, prior systemic treatment of the current cancer, other malignancies, cardiac disease or history of cardiac dysfunction, pregnancy, lactation, and refusal to use contraception.

### Procedures

SBRT was performed every other day in breast target lesion with a total radiation dose of 24Gy separated by 3 fractions. Within 2-4 weeks after the completion of radiotherapy, individual patients received 6 cycles of combination of oral 150 mg Dalpiciclib daily on days 1-21 of each 4-week cycle and 25 mg exemestane with a total duration of 24 weeks. Patients without menopause were also injected subcutaneously with 3.6 mg goserelin every 4 weeks. All patients received study treatment until completion of all prescribed protocol therapy, disease progression, unacceptable toxicity, or withdrawal of consent. If disease progression, the patient was either proceed to surgery or received alternative neoadjuvant therapy. Surgery was performed 3-5 weeks after the last dose of dalpiciclib to allow adverse event (AE) recovery. Recommended surgery and adjuvant therapy followed per local guidelines or institutional standards. The necessity of additional chemotherapy for patients with pCR was judged by investigators. A 5-year follow-up is conducted every 3 months during the first 2 years, and every 6 months for the subsequent 3 years. Additionally, safety data are collected within 90 days after surgery for subjects who discontinue study treatment.

### Outcomes

The primary endpoint was the rate of residual cancer burden (RCB) 0-I. Secondary endpoints included ORR (defined as the proportion of patients with complete or partial response according to the Response Evaluation Criteria In Solid Tumors [RECIST] version 1.1 by MRI), pCR rate in the breast and axillary lymph nodes (tpCR; ypT0/is ypN0) and in the breast (bpCR; ypT0/is ypNx), breast-conservation rate, preoperative endocrine prognostic index (PEPI) score for breast cancer-specific survival (BCSS) (PEPI 0), and safety, according to the Common Terminology Criteria for Adverse Events (CTCAE) version 5.0.

### Patient-Reported Outcome (PRO) assessments

In the study, the patient-reported outcome (PRO) assessments was measured using the European Organization for Research and Treatment of Cancer (EORTC) Quality of Life Questionnaire Core-30 (QLQ-C30; version 3) (Cull, 1998) at baseline, at the end of cycles 1 to 6, and before surgery. The PRO data were scored, according to the EORTC scoring manual. A raw score was calculated as the average of items contributing to a scale and standardized to a range of 0 to 100 points. On global health status and functioning scales, a higher score indicated better status. Higher scores on symptom scales indicated worse symptoms.

### Tumor-infiltrating lymphocytes (TILs)

Tumor samples were collected when performing needle biopsy and surgery. The specimens were fixed in 10% formalin and paraffin-embedded. The tissue sections (3 µm) were stained with hematoxylin and eosin (H&E) and the frequency of TILs were quantified manually by a pathologist in a blinded manner. The TILs scores were determined as the percentage of TILs in tumor area (tumor cells and stroma). The percentage of stromal TILs (sTILs) was calculated in accordance with the recommendations by an International TILs Working Group 2014 (Salgado et al., 2015).

### Programmed cell death-ligand 1 (PD-L1) expression

The expression of programmed cell death-ligand 1 (PD-L1) was characterized by immunohistochemistry (IHC) using anti-PD-L1 (clone SP142, Spring Bioscience, USA) and the percentages of PD-L1 positive tumor cells (TC) or infiltrating inflammatory cells (IC) were quantified.

### Statistical analysis

This exploratory study involves 12 patients, with the sample size determined based on clinical considerations, not statistical factors.

All statistical analyses were conducted using SAS 9.4 (North Carolina, USA). Continuous data are presented as mean and standard deviation (SD) or mean and 95% confidence interval (CI). Categorical data are expressed as frequency and percentage. The 95% CIs of pathological complete response rate, proportion of patients with RCB-0 or RCB-I, and ORR were estimated using the Clopper-Pearson method.

## Results

### Patient characteristics

Between November 2021 and January 2022, 12 HR-positive, HER2-negative patients were screened and recruited in the trial. Their baseline demographic and clinical characteristics are shown in **Table 1**. They had a median age of 48.5 years (range, 36-56) and the median tumor size of 33.5mm (range, 23-90) assessed by breast MRI.

**Table 1:** Baseline characteristics of the patients.

|  | **Patients (n=12)** |
| --- | --- |
| **Median age (range), years** | 48.5 (36-56) |
| $\leq 50$ | 6 (50.0%) |
| $> 50$ | 6 (50.0%) |
| **Menopausal status** |  |
| Premenopausal | 6 (50.0%) |
| Postmenopausal | 6 (50.0%) |
| **Tumor size** |  |
| T2 | 11 (91.7%) |
| T3 | 1 (8.3%) |
| **Lymph node status** |  |
| N0 | 5 (41.7%) |
| N1 | 2 (16.7%) |
| N2 | 5 (41.7%) |
| **Clinical stage** |  |
| IIA | 4 (33.3%) |
| IIB | 3 (25.0%) |
| IIIA | 5 (41.7%) |
| **Tumor grade** |  |
| II | 11 (91.7%) |
| III | 1 (8.3%) |
| **ER expression** |  |
| $> 10\%$ | 12 (100.0%) |
| **PgR expression** |  |
| $< 20\%$ | 3 (25.0%) |
| $\geq 20\%$ | 9 (75.0%) |
| **HER2 expression** |  |
| 0 | 7 (58.3%) |
| 1+ | 1 (8.3%) |
| 2+, FISH | 4 (33.3%) |
| **Median baseline Ki67 (range), %** | 35 (15-70) |
| $\leq 14\%$ | 3 (25.0%) |
| $> 14\%$ | 9 (75.0%) |

Data are n (%) or median (range).
ER: estrogen receptor; PgR: progesterone receptor; HER2: human epidermal growth factor receptor 2; FISH: fluorescence in situ hybridization.

### Patient outcomes

After the completion of study treatment, 2 (16.7%, 95% CI 3.5%-46.0%) out of 12 patients achieved RCB 0–I tumor response, tpCR and PEPI 0 (**Table 2**). There were 3 (25.0%) patients reached bpCR. Overall, this study led to an ORR (as assessed by breast MRI) of 91.7% (95% CI 62.5%-100%, **Fig. 1**). Rate of conversion from mastectomy at baseline to breast conservation at surgery was 25.0% (95% CI 8.3%-53.9%). The median Ki67 of surgical specimens for patients without pCR was 7% (range, 5-70).

**Figure 1:**
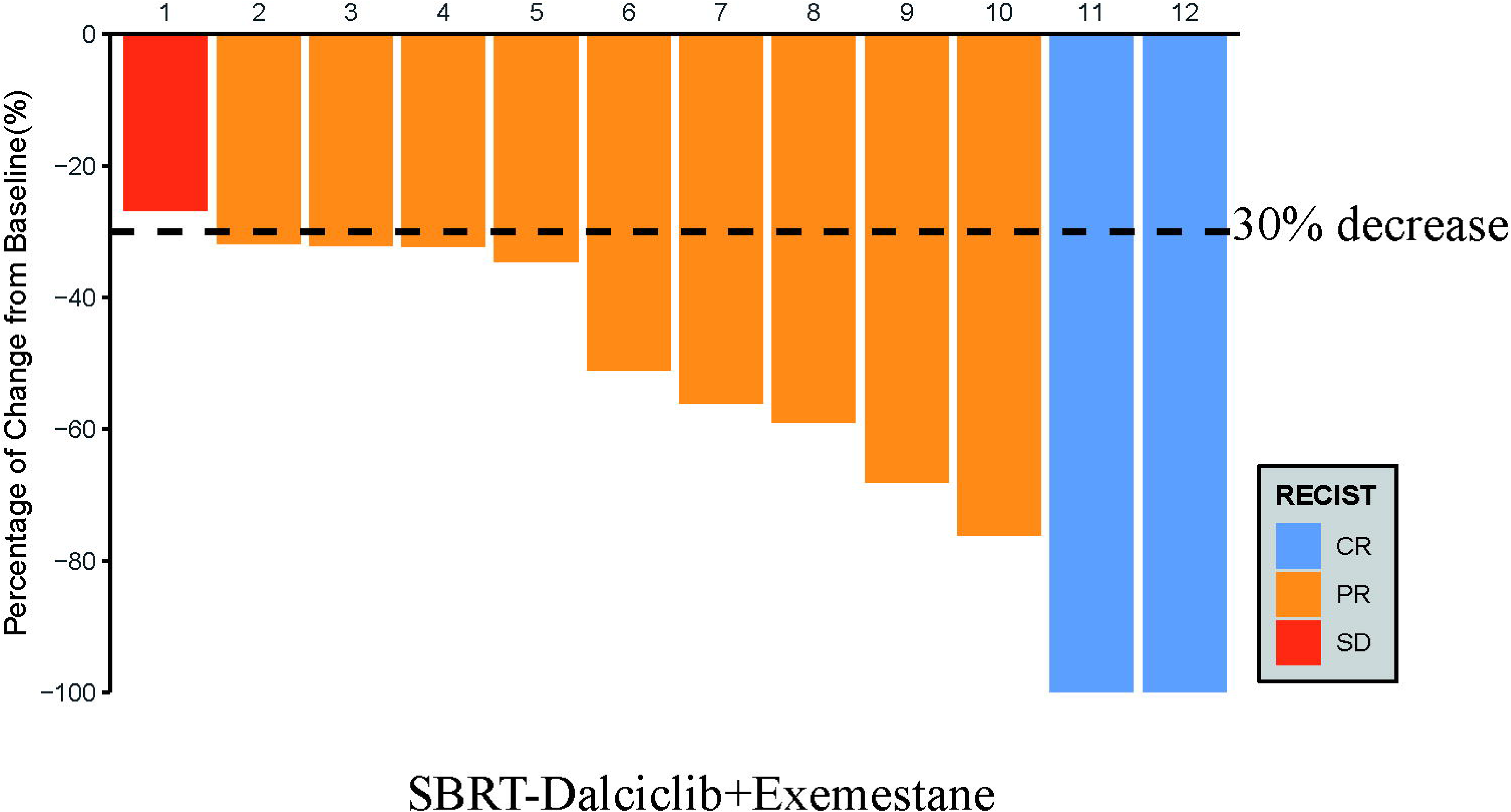
Waterfall plot of best reduction in tumor measurement from basline.

**Table 2:** Pathological and clinical response (n=12).

|  | n (%) |
| --- | --- |
| **Total pathological complete response** | 2 (16.7%) |
| **Breast pathological complete response** | 3 (25.0%) |
| **Residual cancer burden score** |  |
| 0 | 2 (16.7%) |
| I | 0 |
| II | 6 (50.0%) |
| III | 4 (33.3%) |
| **Radiological response** |  |
| Complete response | 2 (16.7%) |
| Partial response | 9 (75.0%) |
| Stable disease | 1 (8.3%) |
| Objective response rate | 11 (91.7%) |
| **Preoperative endocrine prognostic index** |  |
| 0 | 2 (16.7%) |
| 1-3 | 0 |
| ≥4 | 10 (83.3%) |

Data are presented as n (%)

### Toxicity

All patients completed the study treatment and AEs are summarized in **Table 3**. The most frequent grade 1-2 AEs were leukopenia (58.3%), hot flushes (50.0%) and lymphopenia (50.0%). There were some patients with Grade 3 AEs, including neutropenia (66.7%) and leukopenia (25.0%), but not grade 4 AEs. There was no serious AEs, no therapy-related death and no patients with dose interruption and reduction or treatment discontinuation in this population. Furthermore, there was no patient with radiation-related dermatitis and skin hyperpigmentation in this population.

**Table 3:** Treatment-emergent adverse events (n=12).

|  | n (%) |  |  |
| --- | --- | --- | --- |
|  | Grade 1 or 2 | Grade 3 | Grade 4 |
| Neutropenia | 3 (25.0%) | 8 (66.7%) | 0 |
| Leukopenia | 7 (58.3%) | 3 (25.0%) | 0 |
| Hot flushes | 6 (50.0%) | 0 | 0 |
| Lymphopenia | 6 (50.0%) | 0 | 0 |
| Insomnia | 4 (33.3%) | 0 | 0 |
| Alopecia | 4 (33.3%) | 0 | 0 |
| Anemia | 4 (33.3%) | 0 | 0 |
| Hyperuricemia | 4 (33.3%) | 0 | 0 |
| Blood urea increased | 4 (33.3%) | 0 | 0 |
| Rash | 3 (25.0%) | 0 | 0 |
| Thrombocytopenia | 3 (25.0%) | 0 | 0 |
| Hyperglycemia | 3 (25.0%) | 0 | 0 |
| Creatinine increased | 3 (25.0%) | 0 | 0 |
| Fatigue | 3 (25.0%) | 0 | 0 |
| Arthralgia | 2 (16.7%) | 0 | 0 |
| γ-glutamyl transferase increased | 2 (16.7%) | 0 | 0 |
| Hypocalcemia | 2 (16.7%) | 0 | 0 |
| Abnormal T wave of<br>electrocardiogram | 2 (16.7%) | 0 | 0 |
| Hyponatremia | 2 (16.7%) | 0 | 0 |
| Lactate dehydrogenase increased | 2 (16.7%) | 0 | 0 |
| Diarrhea | 1 (8.3%) | 0 | 0 |
| Headache | 1 (8.3%) | 0 | 0 |
| Mucosal inflammation Constipation | 1 (8.3%) | 0 | 0 |
| Proteinuria | 1 (8.3%) | 0 | 0 |
| Hypertriglyceridemia | 1 (8.3%) | 0 | 0 |
| Alanine aminotransferase increased | 1 (8.3%) | 0 | 0 |

Data are presented as n (%).

### Patient-reported outcome

All patients completed the EORTC QLQ-C30 and had available results of PRO assessment (**Fig. 2**). There was no clinically significant deterioration in global health status, physical, role, emotional, cognitive and social functioning in this population.

**Figure 2:**
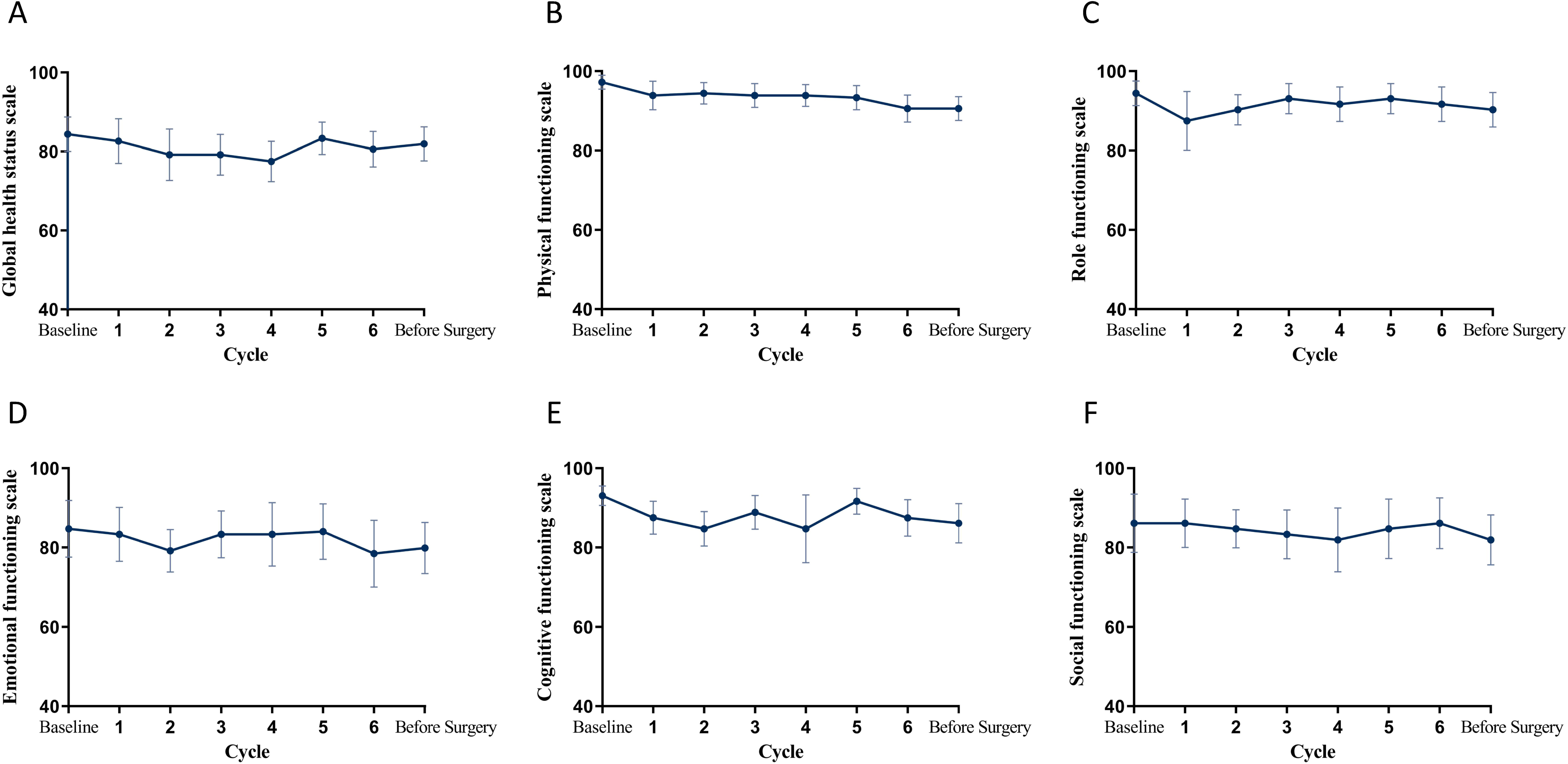
Mean change from baseline in QLQ-C30 over time. (A) Global health status. (B-F) Physical, role, emotional, cognitive and social functioning.

### Alteration in tumor immune microenvironment

To examine the impact of this new neoadjuvant therapy on tumor environment, we performed H&E staining of tissue sections in 10 evaluable patients (**Fig. 3**). Compared with those in pre-treatment biopsied tissues, treatment significantly increased the percentages of tumor-infiltrating lymphocytes (TILs) in the tumor environment (22.9% vs.10.7%, P=0.043). IHC staining of PD-L1 revealed that there appeared no significant difference in the percent of PD-L1 positive immune cells and tumor cells between the pre-treatment and post-treatment specimens.

**Figure 3:**
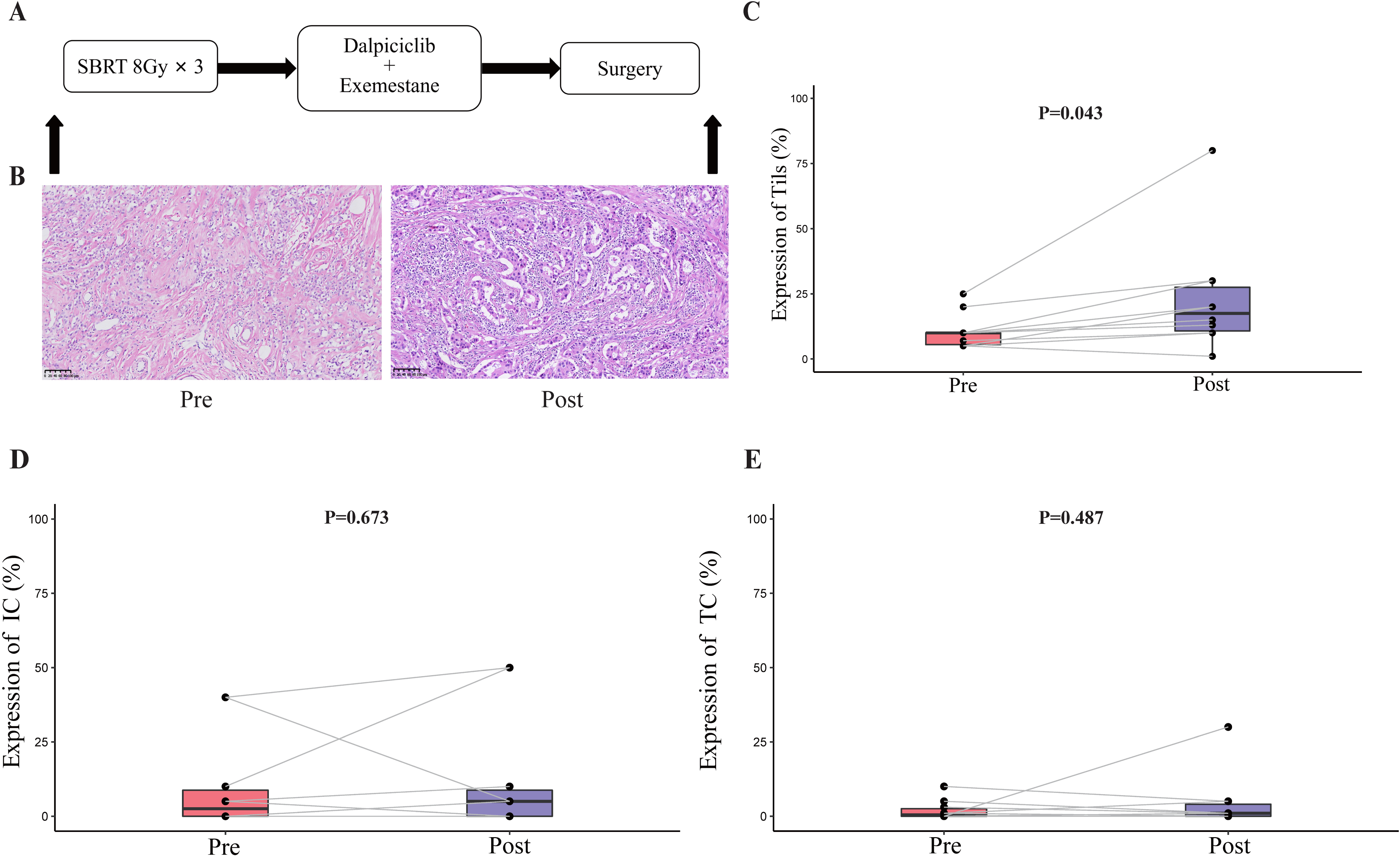
TILs and PD-L1 changes before and after treatment. (A) Study schema. (B) Representative TILs. Scale bar, 100 μM. (C-E) Percentage of TILs, PD-L1 expression in immune cell and tumor cell in paired specimens (n=10). Bars, boxes, and whiskers represent median, interquartile range, and range, respectively.

## Discussion

This pioneering study reported the efficacy and safety of sequential neoadjuvant radiotherapy and combination of dalpiciclib and exemestane in patients with HR-positive, HER2-negative breast cancer. This new therapeutic regimen achieved considerable anti-tumor activity with the RCB 0-I rate of 16.7% and ORR rate of 91.7%, respectively. More importantly, this combinatory therapeutic strategy was well tolerated in this population.

CDK4/6 inhibitors have recently been preclinically identified as radiosensitizers, acting mainly through hindering DNA damage repair response after radiation exposure, and enhancing cell apoptosis and blocking cell cycle, which provide a conceptual basis of combining radiotherapy with a CDK4/6 inhibitor (Khasraw et al., 2017; Xie et al., 2019). Moreover, both radiotherapy and CDK4/6 inhibitors exert immunomodulatory effects and may synergize anticancer immune responses (Andrea M Pesch et al., 2020). SBRT with hypofractionation and higher radiation doses is more effective than conventional fractionation in triggering immune responses (Gandhi et al., 2015). Consistent with the basic research (Giulia Petroni et al., 2021), we used the sequential treatment with SBRT and combination of dalpiciclib and exemestane.

Unlike HER2-positive and triple-negative breast cancer, NCT or NET for HR-positive, HER2-negative breast cancer only achieves a pCR rate of ≤ 10% (Cortazar et al., 2014). Our study indicated that this novel therapeutic regiment achieved a pCR and RCB 0-I rate of 16.7%, while neoadjuvant therapies with combination of a CDK4/6 inhibitor and letrozole led to a pCR rate of 0-3.8% and RCB 0-I rate of 6.1-7.7% and treatment with NCT resulted in a pCR rate of 5.8-5.9% and RCB 0-I rate of 11.8-15.7% in HR-positive, HER2-negative breast cancer patients (C. X. Ma et al., 2017; Prat et al., 2020). It is notable that pCR rate (commonly defined as ypT0/isN0) is not significantly associated with the prognosis of HR-positive, HER2-negative breast cancer (Cynthia X Ma et al., 2017). In addition, clinical trials to test different therapies for patients with HR-positive, HER2-negative breast cancer have focused on clinical response rates as important clinical endpoints (Hurvitz et al., 2020; Johnston et al., 2019; Khan et al., 2020; Khasraw et al., 2017; C. X. Ma et al., 2017). In our study, treatment with the new therapeutic regiment led to eleven (91.7%) patients achieving objective responses by MRI, while previous studies have revealed that NET achieves an objective response rate of approximately 60% in HR-positive, HER2-negative breast cancer patients (Sella et al., 2021). One of the principal objectives of neoadjuvant therapy is to facilitate downstaging and enhance the rate of breast conservation. With the addition of SBRT, a slightly higher rate of patients changed from mastectomy to breast conservation compared with PALLET study (25.0% vs 14.1%). (Johnston et al., 2019). Consequently, our new neoadjuvant therapeutic regiment appeared to improve the likelihood of pathological downstaging and achieve a margin-free resection, particularly for those with locally advanced and high-risk breast cancer.

Considering that treatment with radiotherapy or a CDK4/6 inhibitor can enhance anti-tumor immunity in vitro, We analyzed the impact of new neoadjuvant therapeutic regiment on alternation in biomarker expression in the numbers of TILs before and after the treatment., Our study showed a modest but statistically significant increase in the tumor tissues with the addition of SBRT. Apparently, sequential neoadjuvant therapies with radiotherapy, a CDK 4/6 inhibitor and exemestane turned non-responsive ‘cold’ tumors into responsive ‘hot’ ones. Therefore, our findings may provide significant insights into designing effective therapeutic strategies against HR-positive, HER2-negative breast cancer.

Patients with HR-positive, HER2-negative breast cancer were well-tolerated to the combinational regimen. During the observation period, those patients only developed grade 1-3 of AEs, including grade 3 neutropenia (66.7%), similar to that of combination of dalpiciclib and fulvestrant in the DAWNA-1 trial (84.2%) (Xu et al., 2021). There was no new AEs following therapies with radiotherapy and dalpiciclib in this population.

We recognized that our study had limitations, including a one-arm preliminary exploratory trial with small sample size. The small sample size limited the comparison of our data with historical data in the literature due to the potential bias. Consequently, cross-trial comparisons like this hold limited significance. Thus, further prospective randomized clinical trials with a larger population for longer treatment duration are warranted to validate the findings.

In conclusion, our study provides novel insights into the neoadjuvant treatment of HR+/HER2-breast cancer. The sequential radiotherapy and a CDK4/6 inhibitor plus ET were effective and well-tolerated, which could serve as a feasible option for neoadjuvant therapy for HR-positive, HER2-negative breast cancers. Further validation of these findings is warranted in a large-scale study.

## Acknowledgments

Jiangsu Hengrui Pharmaceuticals provided the study drug dalpiciclib free of charge for patients enrolled in the study. We thank the patients, their families involved in this study.

## Data availability statement

The raw clinical and imaging data are protected due to patient privacy laws. The datasets generated and/or analyzed during the current study are available from the corresponding author Caigang Liu on request for 10 years; de-identified clinical data and experimental data are available on request sharing, which will need the approval of the Institutional Ethical Committees. De-identified data will then be transferred to the inquiring investigator over secure file transfer.

